# Pulmonary Thromboembolic Disease in Patients with COVID-19 Undergoing Computed Tomography Pulmonary Angiography (CTPA): Incidence and Relationship with Pulmonary Parenchymal Abnormalities

**DOI:** 10.1101/2020.06.01.20118505

**Authors:** Cheng Fang, Giorgio Garzillo, Bhavna Batohi, James T H Teo, Marko Berovic, Paul S Sidhu, Hasti Robbie

## Abstract

**Purpose:** This study aims to report the incidence, severity and extent of pulmonary thromboembolic disease (PTD) in patients with confirmed COVID-19 who have undergone CT pulmonary angiography (CTPA) in a tertiary centre.

**Materials and Methods:** This is a retrospective analysis of all patients undergoing CTPA between 23^rd^ March 2020 and 19^th^ April 2020 in a tertiary centre. The presence of PTD, location and involved pulmonary lobes were documented. The pattern and extent of pulmonary parenchymal abnormalities including the presence of fibrosis, lymph node enlargement and pleural effusion were evaluated by two experienced observers independently and consensus was achieved for the most disparate results. Inter-observer agreement was assessed using Kappa statistics. Student t-test, Chi square and Mann-Whitney U tests were used to compare imaging features between PTD and non-PTD sub-groups.

**Results:** During the study period, 2157 patients were confirmed with COVID-19, 297/2157 (13.8%) had CT imaging, 100/2157 (4.6%) were CTPA studies, 93 studies were analysed, excluding suboptimal studies. Overall incidence of PTD was 41/93 (44%) with a third of patients showing segmental and subsegmental PTD (n = 28/93, 30%,). D-dimer was elevated in 90/93 (96.8%) of cases. High Wells’ score did not differentiate between PE and non-PE groups (p = 0.801). The inter-observer agreement was fair (Kappa = 0.659) for parenchymal pattern and excellent (Kappa = 0.816) for severity. Lymph node enlargement was found in 34/93 of cases (36.6%) with 29/34 (85.3%) showing no additional source of infection. Fibrosis was seen in 16/93 (17.2%) of cases, mainly demonstrating fibrotic organising pneumonia.

**Conclusion:** There is a high incidence of PTD in COVID-19 patients undergoing CTPA, complicated by lack of a valid risk stratification tool. Our data indicates a much higher suspicion of PTD is needed in severe COVID-19 patients. The concomitant presence of fibrotic features on CT indicates the need for follow-up for evaluation of chronic pulmonary complications.

## INTRODUCTION

Coronavirus disease 2019 (COVID-19), the pandemic caused by the novel coronavirus known as severe acute respiratory syndrome coronavirus 2 (SARS-CoV-2), was first identified in Wuhan, China in December 2019 [1]. Since then, the virus has spread rapidly worldwide and as of 8^th^ May 2020, there are over 3.7 million reported cases of COVID-19 globally with over 260,000 deaths [2]. The majority of patients with COVID-19 present with an acute respiratory illness of varying severity with fever, cough and dyspnoea being the most common symptoms [3,4].

The diagnosis of COVID-19 is usually confirmed by identification of viral RNA using reverse-transcription polymerase chain reaction (RT-PCR). However, the RT-PCR test is subject to variability depending on sampling technique with positive detection rate ranging between 30–60% [5]. Hence, imaging has become an important tool in the diagnosis and management of patients with COVID-19, initially prompted by findings of a study by Ai T et al [6] confirming high sensitivity of computed tomography (CT) in diagnosing COVID-19. However, the use of CT as first line imaging technique remains controversial and in most radiology departments, CT scanning is reserved for cases with severe disease or those with stagnating or deteriorating clinical conditions [7,8]. In the wake of the pandemic, various radiological societies and colleges have set guidance on how to interpret the pattern and extent of radiographic and CT findings in patients with suspected or confirmed COVID-19 [9–12]. However, none of the existing guidance has been externally validated in larger cohorts. In addition, despite several reports investigating the relationship between poor outcomes and various clinical and biochemical factors [3,4,13,14], the focus on radiological parameters remains somewhat limited.

Coagulopathy is emerging as one of the complications of COVID-19 and several reports have concluded that high D-dimer values, prolonged prothrombin time and thrombocytopenia are associated with poor outcome [3,14–19]. The pathophysiology is not well understood but emerging evidence points towards hyperinflammation resulting in a vascular disease within the lungs that is primarily characterised by thrombotic microangiopathy [19,20]. Recently published data in a small cohort of hospitalised patients with severe COVID-19 suggests up to 40% incidence of pulmonary thromboembolic disease (PTD) [21,22]. The recent report from the national institute for Public Health of the Netherlands concluded that thrombotic state can occur in a substantial percentage of COVID-19 patients but highlighted the paucity of data on the prevalence of PTD [23].

In this study, we aim to report the incidence as well as severity and extent of PTD in patients with confirmed COVID-19 who have undergone CTPA imaging in a tertiary centre. The secondary aim of this study is 1; to assess the effectiveness of British Society of Thoracic Imaging (BSTI) current guidance on pattern and extent of lung parenchymal abnormalities in COVID-19 [11] and 2; to assess whether there is any difference in the patterns and severity of disease on CT in COVID-19 patients with and without PTD.

## MATERIAL & METHODS

### Study population

A single centre retrospective analysis of all consecutive patients with confirmed SARS-CoV-2 on RT-PCR who underwent a CT pulmonary angiography (CTPA) study between 23rd March 2020 and 19th April 2020. This project operated under London South East Research Ethics committee (reference 18/LO/20448) approval granted to King’s Electronic Records Research Interface (KERRI) specific work on COVID-19 research, was reviewed with expert patient input on a virtual committee with Caldicott Guardian oversight. Patient consent was not required due to retrospective nature of the study.

### CTPA protocol

All CT examinations were acquired using a GE Discover 750 HD (64 slice) scanner (GE Healthcare) using our departmental COVID-19 CTPA protocol for PTD (slice thickness 1.25mm, peak tube voltage of 100kVp and current modulation range between 80–500mAs) which includes a volumetric non-contrast high-resolution CT thorax prior to administration of 60mls of intravenous non-ionic contrast with 100mls of a saline chaser at a rate of 4.5 ml/sec with a time delay of 6 sec, using bolus tracking technique. All patients were scanned in a supine position from lung apices to bases at full inspiration. All scans were reconstructed using a high-spatial-frequency soft kernel (WW/WL 400/40, 0.625mm slices). Contiguous images of 1-mm thickness were reviewed on lung window settings for parenchymal disease (width,1500HU; level, −500 HU) and at mediastinal window settings for PTD and lymph node enlargement (width, 350HU; level, 50HU).

### Data collection

Patient demographic, clinical, laboratory and outcome data were extracted from electronic medical records. Laboratory data collected were from the date closest to the CTPA study date. Duration of clinical symptom was defined from the self-reported onset of COVID-19 symptoms to the date of CTPA. Wells’ scores for PTD were calculated [24]. Outcome data was updated until 3^rd^ May 2020 which was two weeks after the end of study inclusion period.

### Image analysis

All CT studies were reviewed by two radiologists (CF with 4 years’ experience in cancer imaging and HR with 6 years’ experience in thoracic imaging). CT pattern of lung disease and severity of parenchyma abnormality, severity and extent of PTD, presence of right heart strain, fibrotic features and intrathoracic lymph node enlargement were reviewed. A consensus was reached by a third radiologist (MB, 11 years’ experience in thoracic imaging) where there was a disagreement. All readers were blinded to patient clinical details and existing radiology reports. CT pattern and the extent of parenchymal disease was documented as normal, classic, probable, indeterminate or non-COVID-19 and mild, moderate and severe respectively based on BSTI guidelines [11] (**Table 1**). The CT severity was defined as mild if up to 3 focal abnormality of 3cm in maximum diameter. Severity of PTD was classified as subsegmental, segmental, lobar, main and saddle. The extent of PTD was documented based on the number of pulmonary lobes involved (between 1 – 6); with lingula considered a separate lobe. Fibrosis was considered present if two of the following were observed: traction bronchiectasis within the areas of parenchymal abnormality, volume loss and architectural distortion as per Fleischner Society glossary of terms [25]. The patterns of pulmonary fibrosis on CT were categorised into fibrotic organising pneumonia or acute respiratory distress syndrome (ARDS), also based on Fleischner Society glossary of terms [25]. Intrathoracic lymph nodes were considered enlarged if they measured > 10mm or > 3mm in short axis diameter for mediastinal and hilar lymph nodes respectively [26]. Right heart strain on CT was defined as pulmonary artery diameter to ascending thoracic aorta ratio > 1 [27].

**Table 1.** Various CT patterns that could represent, raise suspicion of or suggest alternative diagnosis.

| CT Patterns | CT findings |
| --- | --- |
| Classic COVID-19 Pneumonia | Peripheral ground-glass opacities (GGO)<br>Possible crazy paving pattern*<br>Organizing pneumonia (OP)** |
| Probable COVID-19 Pneumonia | Mixture of bronchocentric and peripheral consolidation predominantly in the lower zones<br>Organizing pneumonia<br>GGO scarce |
| Indeterminate | Does not fit in the three other categories with diffuse/patchy GGO that are not peripheral, fibrosis with ground glass, complex CT patterns<br>Similar patterns to the above but clinical context is wrong or suggestive of alternative diagnosis |
| Non-COVID | Lobar pneumonia<br>Cavitating consolidation<br>Tree-in-bud/centrilobular opacities, lymph node enlargement, pleural effusions |

for COVID-19 pneumonia as per BSTI guidance.
\*Crazy paving pattern on CT includes GGO with inter/intralobular septal thickening
\*\*Organizing Pneumonia patterns on CT are bronchocentric/peripheral consolidation with reversed halo sign and peri-lobular pattern (25).

### Statistical analysis

Statistical analysis was performed by using SPSS software (IBM, Chicago, version 23); P values < .05 were considered to indicate statistical significance. Continuous variables were reported as mean ±standard deviation or median (interquartile range) as appropriate. Kappa statistics were used to assess inter-observer agreement with the level of agreement classified as: poor (kappa < 0.40), fair (0.4<kappa< 0.6), good (0.6<kappa< 0.75), excellent (0.75<kappa< 1.0) [28]. Student t test, Chi-square test, Mann-Whitney U test where appropriate were used to compare study parameters between the groups.

## RESULTS

### Patent demographics, clinical, laboratory data within PTD and non-PTD subgroups

There were 2157 patients with positive RT-PCR test for SARS-CoV-2 during the study period (**Figure 1)**. Out of 297 patients referred for CT imaging, 100 CTPA studies were performed in 97 COVID-19 patients. Seven studies were excluded from final analysis due to non-diagnostic quality. CTPAs were referred from the emergency department (n = 24/93), inpatient wards (n = 49/93) and intensive care units (n = 20/93). Patient demographic, clinical and laboratory and outcome data are listed in **Table 2**. D-dimer was elevated in 90/93 (96.8%) of study cases. The median duration of symptoms to CTPA studies in patients referred from emergency department, inpatient wards, intensive care unit was 8.5 days, 16 days and 21 days respectively and the overall median duration of symptoms to CTPA studies was 14 days. The overall incidence of PTD was 44% (n = 41/93). In 12/24 (50%) referrals from the emergency department the studies were positive for PTD, 16/49 (32.7%) from inpatient wards and 13/20 (65.0%) from intensive care. In 28/41 (68.3%) of cases there were segmental and sub segmental PTD with 26/41 (63.4%) having three lobes or fewer affected by pulmonary emboli.

**Table 2.** Demographic, clinical and laboratory findings from the patients at the time of CTPA study.

| **Presence of Pulmonary Thromboembolic Disease** |  |  |  |
| --- | --- | --- | --- |
|  | Yes | No | Unadjusted, p-value |
| Number of patients | 41 | 52 |  |
| Parameter |  |  |  |
| **Age – median (IQR)** | 62 (56-69) | 57 (50.5-68.5) | p=0.044 |
| **Gender – no.** | 41 | 52 | p=0.499 |
| Female | 13 | 20 |  |
| Male | 28 | 32 |  |
| **Current smoker – no.** | 5 | 8 | p=0.660 |
| **Comorbidity – no.** |  |  |  |
| Hypertension | 20 | 24 |  |
| Diabetes mellitus | 14 | 24 |  |
| Chronic obstructive lung disease | 2 | 4 |  |
| Malignancy | 6 | 4 |  |
| Chronic kidney disease | 6 | 4 |  |
| Coronary heart disease | 2 | 7 |  |
| Asthma | 2 | 9 |  |
| Obesity (BMI>30) | 13 | 18 |  |
| None of the above | 7 | 9 |  |
| **Referral Source – no. (%)** |  |  |  |
| Emergency department | 12 (29.3%) | 12 (23.0%) |  |
| Intensive care unit | 13 (31.7%) | 7 (13.5%) |  |
| Inpatient Ward | 16 (39.0%) | 33 (63.5%) |  |
| **COVID-19 symptomatic disease duration prior to CTPA – no. days (IQR)** | 16 (9-23) | 13 (7-17.25) | p=0.246 |
| **Wells scores – no.** |  |  | p=0.801 |
| <4 points | 20 | 24 |  |
| >4 points | 21 | 28 |  |
| **Laboratory findings – median (IQR)** |  |  |  |
| White blood cell count (x10<sup>9</sup> per L) | 10.36 (8.53-13.98) | 7.81 (5.64-10.76) | p=0.058 |
| <4 – no. (%) | 0 (0%) | 4 (7.7%) |  |
| 4-10 – no. (%) | 19 (46.3%) | 31 (59.6%) |  |
| >10 – no. (%) | 22 (53.7%) | 17 (32.7%) |  |
| Lymphocyte count (x10<sup>9</sup> | 1.11 (0.73-1.55) | 1.07 (0.74-1.59) | p=0.670 |
| per L) |  |  |  |
| <0.8 – no. (%) | 13 (31.7%) | 17 (32.7%) |  |
| Neutrophils count (x10<sup>9</sup> per L) | 8.52 (6.48-11.61) | 6.07 (4.31-9.67) | p=0.043 |
| Haemoglobin g/L | 111 (84-126) | 121 (102-134.5) | p=0.071 |
| Platelet count, x10<sup>9</sup> per L | 340 (245-412) | 289 (206-421) | p=0.521 |
| Albumin g/L | 27 (24-33) | 33 (29.5-37) | p=0.001 |
| ALT U/L | 44 (32-62.5) | 45 (27.5-80.5) | p=0.222 |
| Creatinine umol/L | 82 (69-142) | 73 (59.5-126) | p=0.557 |
| Urea | 9.2 (4.8-14.1) | 6.7 (4.2-9.15) | p=0.018 |
| C-reactive protein | 146 (92-216.3) | 73 (59.5-126) | p=0.068 |
| LDH u/L | 466 (389-600.75) | 561.5 (439.75-709.25) | p=0.221 |
| Troponin I | 37 (19.5-99) | 17 (8-34) | p=0.759 |
| D-dimer | 7465 (3835-8000) | 2450 (1170-3985) | p=0.001 |
| Serum Ferritin ug/L | 837.5 (616.25-1371.5) | 1051.5 (678.25-2396.5) | p=0.138 |
| **Outcomes – no. (%)** |  |  |  |
| Death | 6 (14.6%) | 5 (9.6%) | P=0.457 |

p-values were using Student's t-test, Chi square and Mann-Whitney U test respectively.

**Figure 1.**
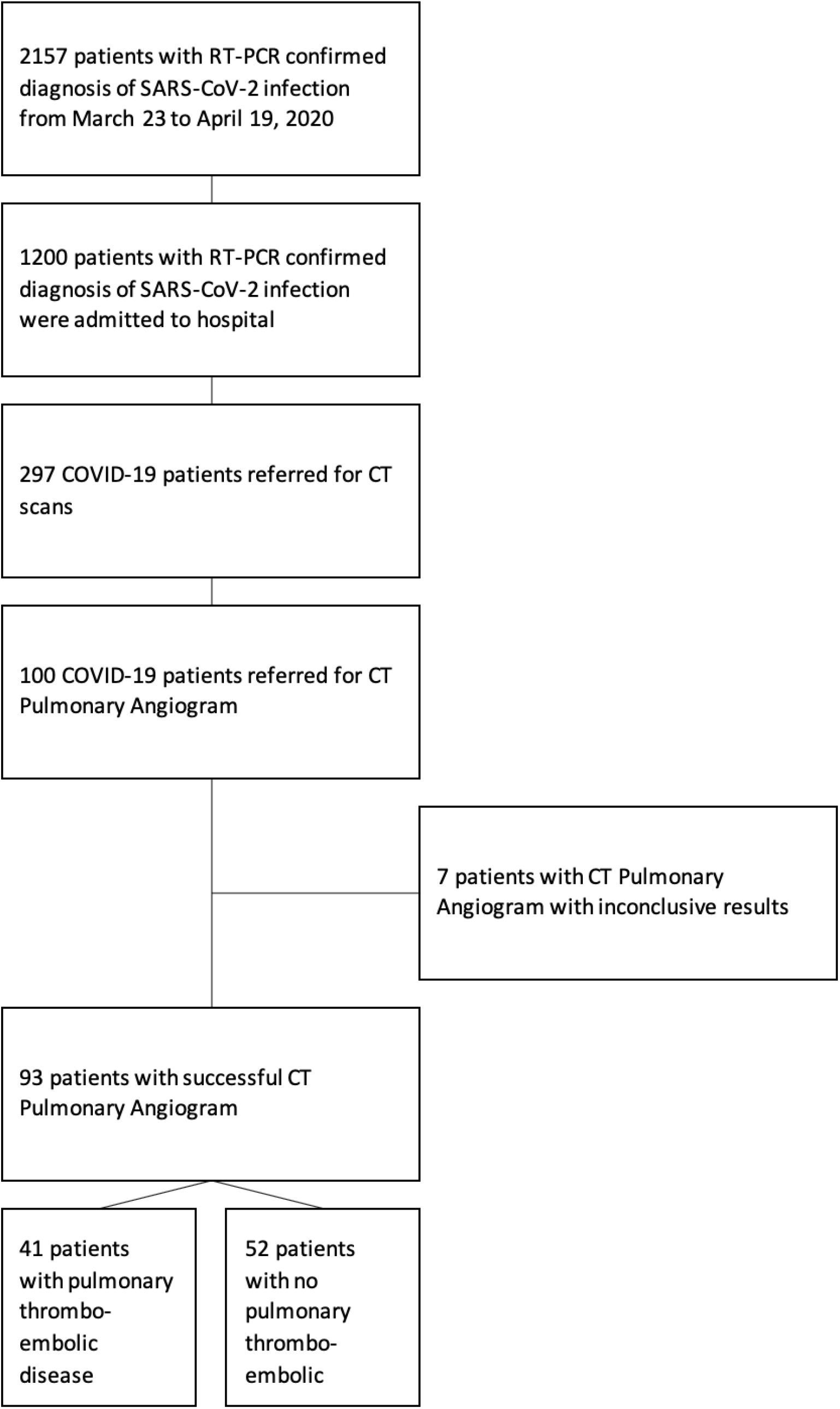
Consort diagram illustrating the process of selecting patients for the final study

### Interobserver agreement

Interobserver agreement for CT pattern of parenchymal disease was fair (Kappa = 0.576, P< 0.001). By grouping the classic and probably categories of lung parenchymal abnormalities, the Kappa increased from fair to good (Kappa: 0.659, P< 0.001). The interobserver agreement was excellent for severity of lung parenchymal changes (Kappa = 0.816, P< 0.001) and fair for pulmonary fibrosis (Kappa = 0.594, P< 0.001)

### CT pattern of the lung disease, nodal status, presence of pleural effusion and extent of pulmonary thromboembolic disease

CT patterns, severity of parenchymal disease and the extent and severity of PTD are detailed in **Table 3**. In 40/93 (43.0%) studies there was a CT pattern that is intermediate for COVID (**Figure 2**). There were 59/93 (63.4%) studies there was severe parenchymal disease with 30/93 (32.2%) showing moderate disease (**Figure 3**). In 16/93 (17.2%) studies imaging demonstrated at least two CT signs of fibrosis. In 13/16 (81.3%) of these cases, the pattern of fibrosis on CT was in keeping with fibrotic organising pneumonia rather than ARDS related fibrosis (**Figure 4**).

**Table 3.**
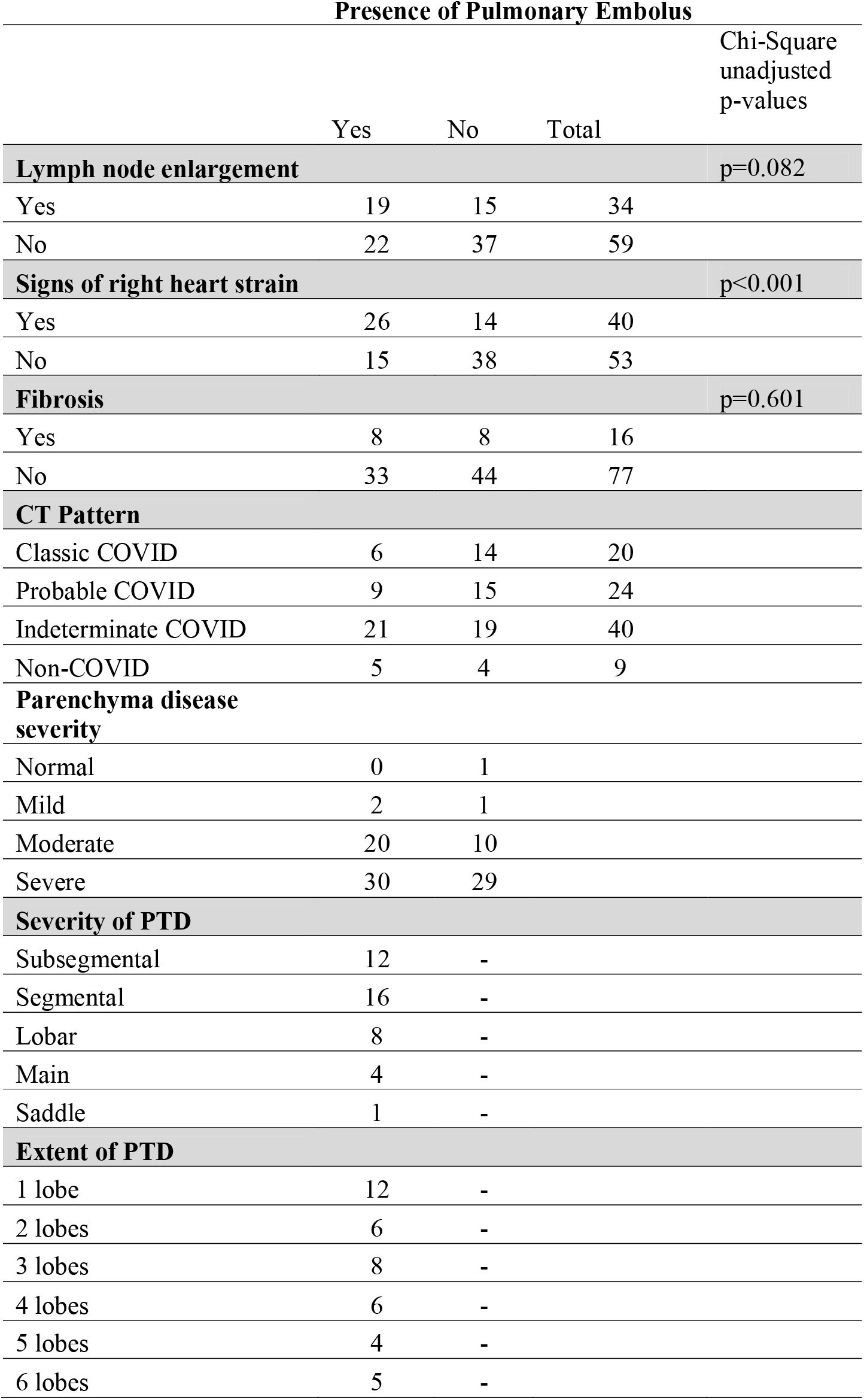
CT patterns in COVID-19 patients who underwent CTPA studies and extent of pulmonary embolism in patients who are found to have positive CTPA studies.

**Figure 2.**
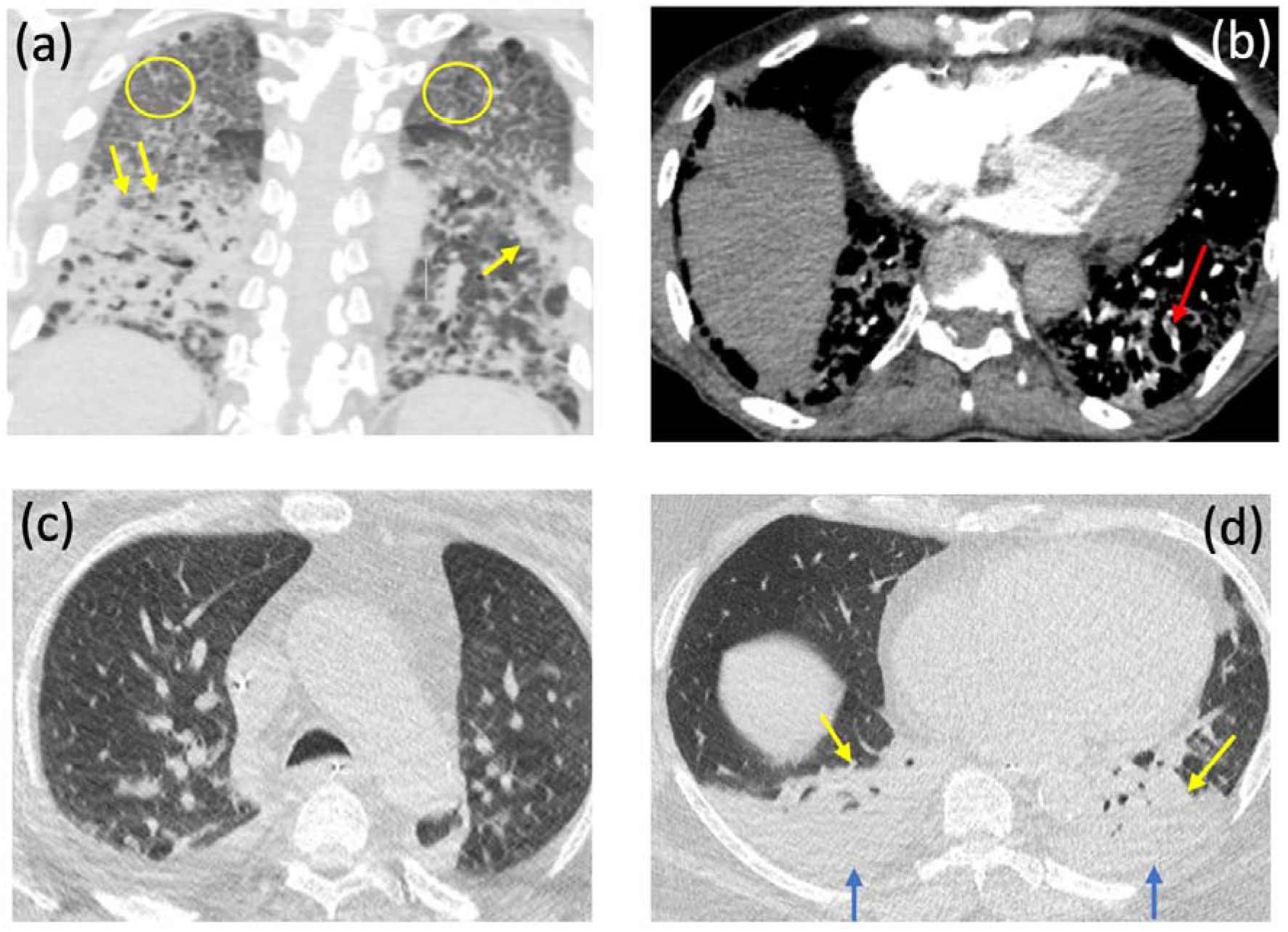
Figures 2a shows axial and coronal non-contrast enhanced CT images of the chest in a 76-year-old male admitted to the ward with covid-19 and increasing O2 requirements. Note bilateral diffuse crazy paving pattern (yellow circles) and lower lobe predominant asymmetrical consolidation (yellow arrows), in keeping with severe disease that is indeterminate for covid. Figure 2b shows the subsequent axial CTPA image confirming small subsegmental PE in the left lower lobe (red arrow). Figures 2 d,e show axial non-contrast enhanced CT images of 67 year old male admitted to intensive care with covid-19 and failure to respond to treatment. Figure 2c shows no parenchymal abnormality in the upper lobes with figure 2d showing bilateral pleural effusions (blue arrows) with lower lobe predominant consolidation and collapse (yellow arrows), in keeping with non-covid pattern.

**Figure 3.**
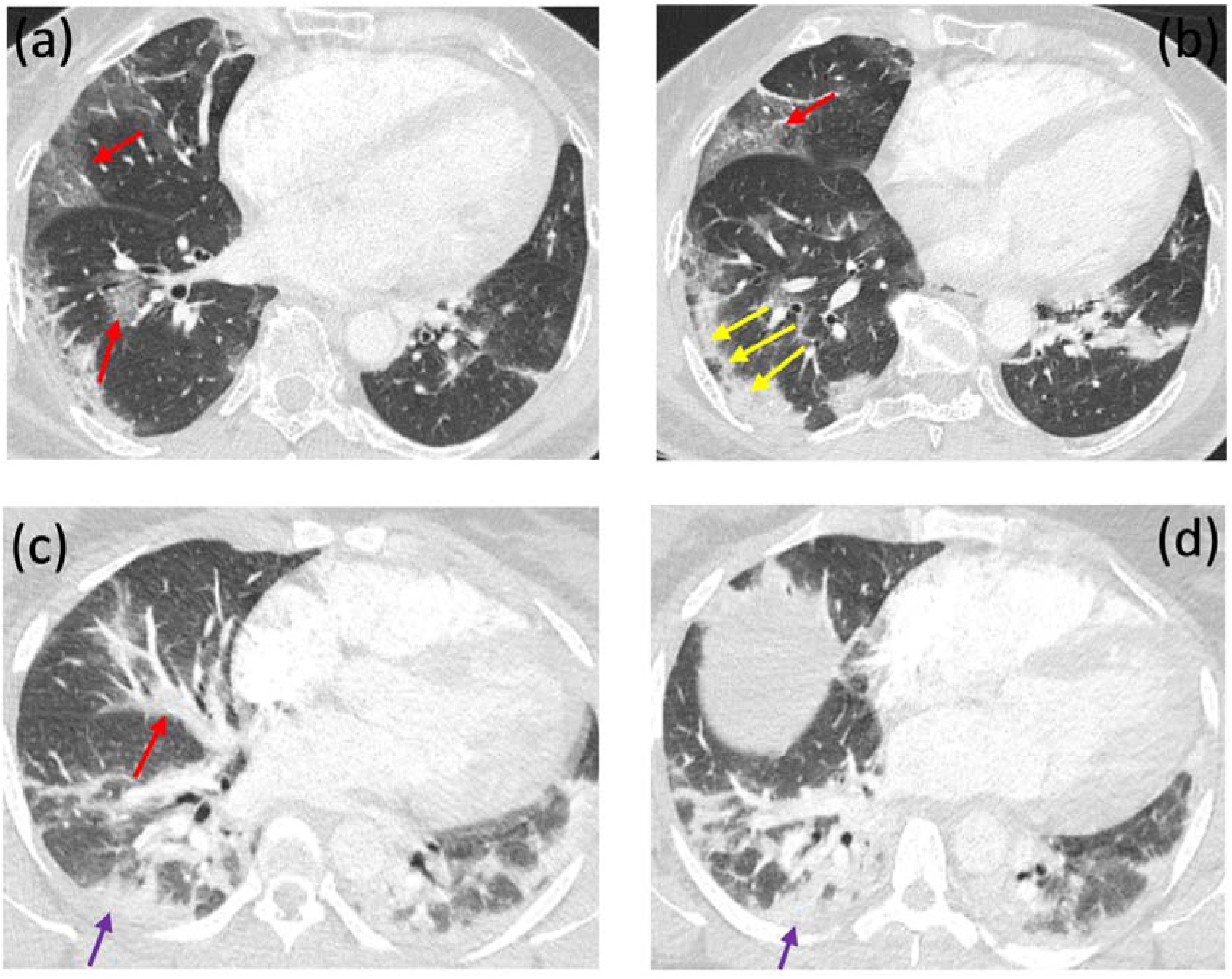
Figures 3 a,b show axial CTPA images of a 36 year old female presenting to emergency department with breathlessness and haemoptysis. Note lower zone predominant peripheral and bronchocentric groundglass opacification (red arrows) and consolidation with a perilobular pattern (yellow arrows). The pattern is in keeping with moderate classic covid-19. Figure 3 d,e show axial CTPA images of a 37 year old female presenting to emergency department with breathlessness. Note bronchocentric (red arrow) and peripheral lower zone consolidation (purple arrows) with no groundglass opacification. The pattern is in keeping with probable covid-19 which is moderate in extent.

**Figure 4.**
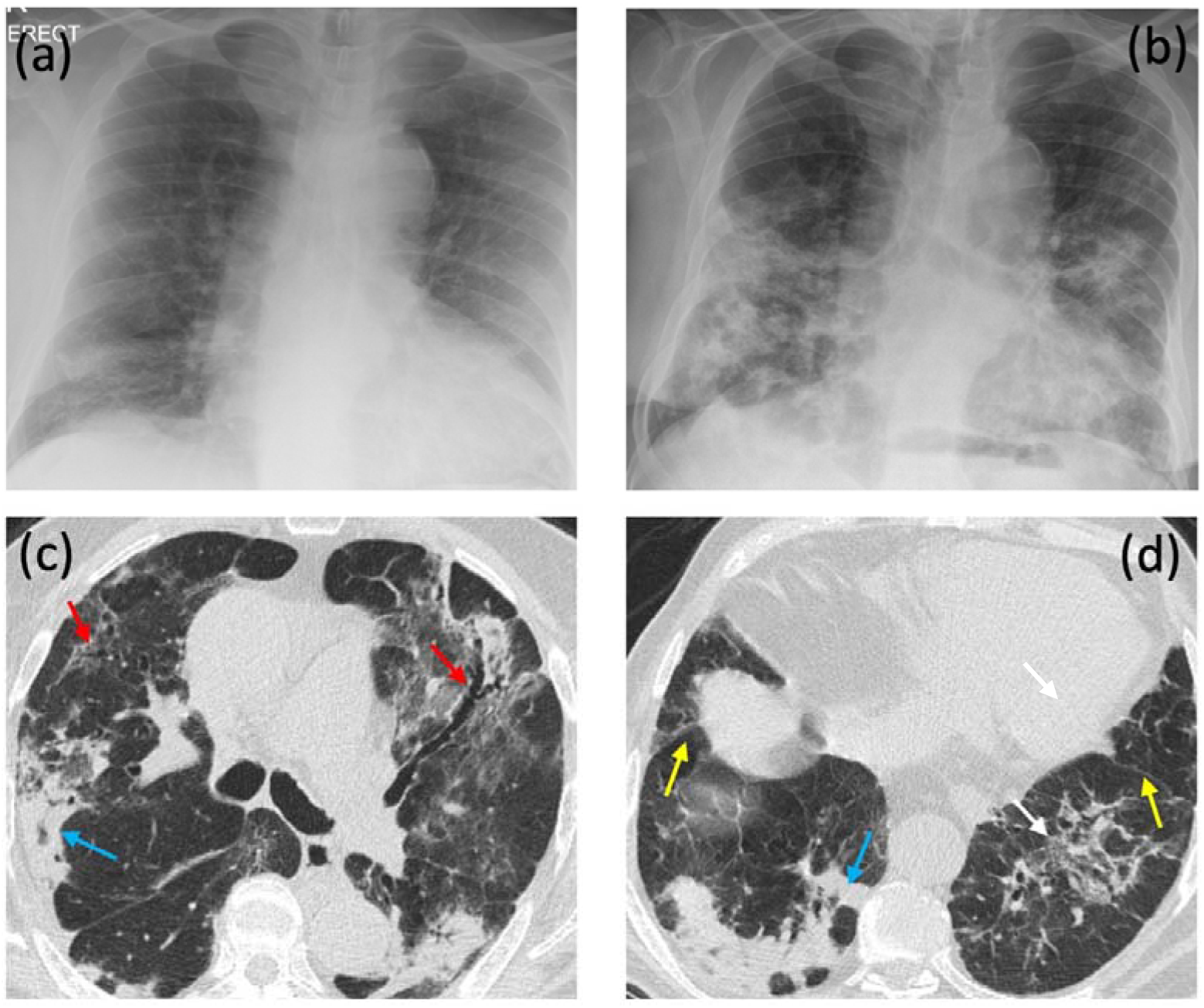
Figures 4 a,b show respective baseline and two week followup chest X-rays of an 84 year old male with history of covid-19, representing to emergency department 4 days post discharge with hypoxia and breathlessness. Note progressive consolidation and architectural distortion in the lower lobes occurring in the interval between the two films. Figures 4 d,e show axial non-contrast CT images of the chest in the same patient. Note peripheral consolidation with peri-lobular pattern (blue arrows) and bronchocentric groundglass opacification with traction bronchiectasis (red arrows), architectural distortion (white arrow) with associated volume loss as seen by posterior retraction of the interlobar fissures (yellow arrows).

There were 34/93 (36.6%) cases with enlarged mediastinal or hilar lymph nodes. The distribution of lymph node enlargement and pleural effusion among different CT patterns is detailed in **Table 4**. The frequency of lymph node enlargement was observed equally amongst different CT patterns, between 30–42.5%. Lymph node enlargement was present in 5/16 (31.3%) of cases who had features of fibrosis versus 29/77 (37.7%) who did not have fibrotic features. Five cases who had lymph node enlargement also had confirmed super-added fungal infection from blood serum with all of these cases showing lymph node enlargement. In the remainder of 29/34 (85.3%) cases with lymph nodes enlargement, there was no additional source of infection. 20/93 (21.5%) of studies in our cohort had pleural effusion.

**Table 4.** The incidence of lymph nodes enlargement and pleural effusion among different CT patterns.

|  | Consensus CT patterns |  |  |  | Total |
| --- | --- | --- | --- | --- | --- |
|  | Classic | Probable | Indeterminate | Non-COVID |  |
| Total | 20 | 24 | 40 | 9 | 93 |
| Lymph node enlargement |  |  |  |  |  |
| No Enlargement | 14 (70%) | 16 (66.7%) | 23 (57.5%) | 6 (66.7%) | 59 |
| Enlargement | 6 (30%) | 8 (33.3%) | 17 (42.5%) | 3 (33.3) | 34 |
| Pleural Effusion |  |  |  |  |  |
| No pleural effusion | 15 (75%) | 24 (100%) | 30 (75%) | 4 (44.4%) | 73 |
| Pleural effusion | 5 (25%) | 0 (0%) | 10 (25%) | 5 (55.6%) | 20 |

The presence of right heart strain was significantly more common among patients with PTD. The level of D-dimer is significantly higher in patients with severe parenchyma disease compared with moderate disease (P = 0.046). The lymph node status and presence of pulmonary fibrosis did not differ among the PTD and non-PTD subgroups **(Table 4)**.

## DISCUSSION

Our study has confirmed the high incidence of PTD in COVID-19 patients who underwent CTPAs. In the pre-COVID-19 period, the detection rate of PTD was 4.2% in the emergency setting without risk stratification and 11.2% with Wells’ score risk stratification [29], whereas, in our COVID-19 cohort, PTD is not only common, but the Wells’ score was not discriminatory in estimating the risk of PTD. This is likely due to Well’s scoring being weighted towards PTD as the leading diagnosis rather than a secondary diagnosis concurrent with symptomatic lung disease.

While conventional D-dimer thresholds was non-discriminatory for PTD (since virtually all COVID-19 patients had elevated D-dimers), we found higher D-dimer levels in COVID-19 patients with PTD versus COVID-19 patients without PTD. This could be explained with higher levels of D-dimer being associated with more severe COVID-19 disease [17].

The radiological pattern and severity of lung disease does not differ in subgroups of patients with or without PTD. The majority of patients in our cohort had segmental and sub-segmental PTD with most patients having three or less lobes affected by thromboembolic disease. Recent pathophysiological data suggests an intravascular inflammatory process occurring in COVID-19 leading to microangiopathic endothelial damage [30]. Our results are somewhat in line with the hypothesis that the vascular disease starts in small vessels within the lungs given that the majority of cases in our cohort had segmental and subsegmental PTD. Virtually all cases developed the PTD in the 2^nd^ to 3^rd^ week of the COVID-19 disease indicating the thromboembolic disease process occurs later during the course of the disease. Our data suggests that the pulmonary thromboembolic event is common in later disease course (2^nd^ and 3^rd^ week of symptomatic disease), but it is currently unknown if thrombosis would also be commonly encountered if a CTPA was performed earlier.

The recommended UK-wide guidance at the time of our study was published by BSTI, which is based on expert consensus rather than real world data. We used the BSTI guidance for characterisation of the CT patterns and the extent of disease on the basis of our current clinical practice as opposed to the more recently introduced reporting systems such as CO-RADS [31]. We found better interobserver agreement using BSTI guidance in our cohort in comparison with reported kappa values of 0.47 for CO-RADS system. However, our results showed that inter-observer agreement for the patterns of parenchymal abnormalities was at best moderate, indicating that BSTI guidance may require revision with more specified definitions for disease patterns. It should also be noted that the majority of cases in our cohort had moderate to severe disease which could mask specific features of COVID-19 with peripheral and lower lobe predominant round ground glass opacities [9] more visible in mild disease. We postulate that this may also be the reason for a relatively large proportion of our patients falling into the indeterminate category because of associated features of diffuse alveolar damage and fibrosis. A proportion of our cohort also demonstrated features suggestive of fibrosis. Although the numbers are relatively small in our cohort, given the number of patients affected globally, this finding indicates that a substantial number of patients with COVID-19 may progress to fibrosing interstitial lung disease, irrespective of the presence of ARDS.

We found a large proportion of patients with COVID-19 to have enlarged intrathoracic lymph nodes which is in distinction to the initial report from China suggesting only 6% incidence of lymph node enlargement in hospitalised patients [32]. Recent correspondence by French investigators [33] showed up to 66% incidence of lymph node enlargement in COVID-19 patients admitted to intensive care unit. This indicates controversy surrounding the issue of lymph node enlargement which could be partly due to differences in study cohorts. We found no co-existing pathogens that could explain lymph node enlargement in the majority of our cases, nor can we attribute the size of the lymph nodes to the presence of fibrosis. Similarly, pleural effusion is regarded as a non-COVID feature however [11], this is also relatively common in our cohort of confirmed COVID-19 cases, but may be a feature of comorbidities.

There are limitations to our study. The sample size is relatively small, and the data was collected retrospectively comprising patients admitted to hospital which result in selection bias of patients with more severe disease.

In conclusion, the high incidence of PTD in COVID-19 patients with severe disease undergoing CTPA, current lack of reliable risk stratification tools and symptoms of PTD overlapping with severe COVID-19 disease, raises the question of whether more CTPA studies should be performed in the 2^nd^ and 3^rd^ week of the illness. Our study suggests that patients with severe disease may require follow-up for long term pulmonary fibrotic complications.

## Data Availability

The dataset used in this will be available on request. The data has not been shred on any other repositories.

